# Patient Perceptions of a Seizure Service Dog in the Epilepsy Monitoring Unit

**DOI:** 10.64898/2026.04.30.26352073

**Authors:** Lia D. Ernst, Bahar Madani, Danny Zhu, Matthew McCaskill, Marissa A. Kellogg

**Affiliations:** Department of Neurology, School of Medicine, Oregon Health & Science University, 3181 Sam Jackson Park Road, Portland, OR 97239, USA; Department of Public Health, Portland State University, Portland, OR 97201, USA; Student Researcher, Department of Neurology, Oregon Health & Science University, 3181 Sam Jackson Park Road, Portland, OR 97239, USA; Portland VA Healthcare System, 3710 SW US Veterans Hospital Rd, Portland, OR 97239

**Author notes:** **Corresponding Author** Lia de Leon Ernst, MD, Department of Neurology Oregon Health & Science University, 3181 Sam Jackson Park Road Portland, OR 97239, USA.

**Keywords:** Seizure dog, seizure detection, service dogs, epilepsy monitoring unit (EMU), seizure safety

## Abstract

**Objective:** Seizure dogs are service animals trained to respond supportively to seizures in people with epilepsy; some are also trained to detect seizure-specific scents, particularly ictal volatile organic compounds (VOCs). This survey study examines feasibility and safety of incorporating a seizure service dog (SSD) into an inpatient setting, as well as patient perceptions of having an SSD in the Epilepsy Monitoring Unit (EMU).

**Methods:** Our SSD underwent specialized training for seizure response and seizure recognition based on seizure-specific VOCs, and accompanied his epileptologist owner in the EMU on rounds for over four years prior to the study. We administered surveys to patients hospitalized in the EMU before and after interactions with a trained seizure dog. The surveys assessed the patients’ comfort with the dog, perceived usefulness of service dogs, safety, and tolerability. Select case examples are also presented in which seizure dog spontaneously alerted prior to epileptic seizures; seizures later confirmed by independent EEG review.

**Results:** Patient responses underscored overall high enthusiasm for seizure dog therapy, with 93% of participants reporting feeling “very comfortable” or “extremely comfortable” with a seizure dog present. No adverse concerns or negative experiences were reported by participants. 91% reported personally experiencing benefits of working with the seizure dog, citing emotional and comfort benefits during their hospitalization. 94% of participants were comfortable with physical contact with the dog or had no proximity preference.

**Conclusion:** These findings suggest that seizure service dogs can be safely integrated into the inpatient EMU setting and have potential to enhance patient care and emotional well-being during EMU monitoring.

**Summary Points:**

- Total of 98 patients admitted to EMU were surveyed about opinions regarding seizure dogs and comfort with integration of seizure dog in EMU setting, with 35 patients completing post-test surveys after interacting with the seizure dog.
- 93% of surveyed EMU patients completing post-test surveys felt very or extremely comfortable with the seizure dog; no negative experiences or safety concerns were reported.
- 91% reported personally experiencing emotional benefits of working with the seizure dog.
- Select case examples demonstrate that the trained seizure dog in our study may be able to spontaneously identify epileptic seizures.

## 1. Introduction

Epilepsy is the fourth most common neurological disorder, affecting one in 26 people over the course of their lifetimes.^1^ People with epilepsy often use dogs for emotional support and assistance during post-seizure recovery. Seizure service dogs (SSD) are specifically trained to assist patients with epilepsy after seizures, and some owners report that their seizure dogs can sense seizures before they begin. There has been interest in utilizing SSD to recognize and/or respond to seizures as a complementary seizure management strategy since the 1990s, when a dog enthusiast magazine published owner reports of dogs being able to recognize seizures.^2^ A subsequent pilot study of six patients and their trained dogs in 1999 suggested that service dogs could in fact be trained to both recognize seizures and provide signals warning their owners within 15 to 45 minutes prior to a seizure occurring.^3^ Two studies surveyed families of people with epilepsy who owned dogs, who self-reported that their dogs exhibited seizure-predictive behaviors in 15% and 10%, respectively, while dogs displayed seizure-responsive behaviors in 40% and 31%.^4,5^

Recent research suggests some dogs can detect seizures by scent, specifically via detection of seizure-specific volatile organic compounds (VOCs). During seizures, people with epilepsy emit specific biomarkers, including unique VOCs associated with physiological changes occurring before, during, and after a seizure. Service dogs can be trained to detect volatile organic compounds (VOCs) associated with ictal sweat. A 2021 study showed that properly trained seizure dogs can detect ictal VOCs in sweat samples with 93.7% accuracy.^6^ Another study in *Scientific Reports* tested five trained dogs in their ability to discriminate seizure odor from non-seizure odor with average sensitivity of 87% and specificity of 98%; three of five dogs had perfect accuracy.^7^

Patients with epilepsy often express enthusiasm about having or acquiring service dogs and have questions about whether they may be able to identify and/or respond to seizures. There is one prior randomized controlled trial investigating benefits of dog ownership for patients with epilepsy. The EPISODE study from the Netherlands randomized patients to standard epilepsy care alone vs. being assigned a seizure dog and investigated seizure outcomes, showing an average 37% reduction in seizure frequency in the seizure dog group, as well as an increase in number of seizure-free days and improved quality of life scores. Seizure dogs offered peace of mind, prevented injuries, and promoted greater independence. They also observed that seizure dogs positively influenced patient cooperation and reduced stigma, underscoring their value beyond seizure detection.^8^

While there are studies supporting benefits of SSD ownership among patients with epilepsy, there is sparse literature regarding potential benefits of integrating seizure dogs in the inpatient epilepsy monitoring unit (EMU) setting. This study aims to broaden understanding of whether it is feasible to utilize service dogs in the EMU setting, how patients feel about the presence of SSD in the EMU, and we also hope to provide preliminary evidence that dogs may be able to accurately identify epileptic seizures by scent in clinical practice.

## 2. Methods

### 2.1 Seizure dog training and background

At our institution, epileptologist Dr. McCaskill has incorporated his seizure dog, Reggie, into patient rounds in the EMU since August 2021. Reggie was initially approved to accompany Dr. McCaskill in the hospital as his personal service dog. When the question was raised by hospital leadership regarding interacting with patients, Dr. McCaskill accepted all liability and provided documentation of Reggie’s training and veterinary care, demonstrating that his seizure dog had greater or equal training to service dogs that were already being utilized as therapy dogs at OHSU’s Doernbecher Children’s Hospital.

Reggie is a golden retriever who has been trained to detect seizure odors and respond to seizures. Reggie has also been trained to respond to seizures by seeking help and providing comfort to patients. Dr. McCaskill incorporates his seizure dog into patient care with consent from patients. We utilized the opportunity to study patient attitudes toward “canine-assisted care” in the inpatient EMU setting and made observations about individual cases when Reggie was present before or during seizures during inpatient care.

The seizure dog’s training was based on the methods employed by the techniques developed by Dr. Jennifer Cattet Ph.D., founder, and principal instructor at the Center for the Study of Medical Assistance Canines.^9^ Methods were adapted for clinical use by training using isolated ictal and interictal sweat samples from people with epilepsy instead of lab-produced isolated VOCs.

Training consisted of virtual video consultations with Dr. Cattet, followed by approximately 8 months of daily 30-60 minute training sessions between the handler and the dog. The dog was trained to tap his nose on the handler’s right thigh repeatedly to indicate presence of seizure scent, after which then the dog will lead the handler to the location of the source of the ictal scent.

### 2.2 Survey protocol

Patients aged 18-89 who were admitted to the EMU for video EEG monitoring in 2023-2024 and had capacity to consent to procedures were considered eligible to participate. Patients who were unable to consent for procedures on their own were excluded from participation. Written consent was obtained, and consented participants completed pre-interaction surveys aimed to assess baseline comfort and attitudes toward the presence of a medical service dog in the EMU. Target enrollment goal was 100 participants. Patients were informed that they may or may not have interactions with the SSD during their admissions.

A baseline survey was provided to consented participants [**Supplement S1: pre-test survey**]. The pre-survey consisted of 15 multiple choice questions with four write-in questions and subject age. If patients ultimately did interact with the seizure dog, they were then asked to complete post-interaction survey [**Supplement S2: post-test survey**]. The post-interaction survey consisted of 10 multiple choice questions and four open-ended questions. Surveys were de-identified and given participant numbers, collected by investigators and stored securely. Data were compiled and analyzed using Microsoft Excel within a secure password-protected database. Incomplete surveys were excluded.

### 2.3 Seizure Examples

Select participants had seizure assessments done by Dr. McCaskill and Reggie the seizure dog. In these select cases, the spontaneous presence of Reggie’s seizure signal was compared to video EEG results by two independent epileptologists to confirm epileptic seizures and verify seizure recognition accuracy.

As part of standard EMU protocol, when patients have seizures, nurses enter the patient’s room to do a seizure assessment, which includes assessment for safety, vital signs and focused physical exam, followed by post-seizure care. Following the nursing assessment, attending physicians often do their own assessments when they are available. At other times, patients may have seizures coincidentally while the attending is in the room. In such cases when Dr. McCaskill was on service in the EMU and he and his service dog were in the patient room, there were instances when the dog would signal during seizures or shortly before seizures. Reggie will indicate to Dr. McCaskill that the scent has been detected by tapping its nose to Dr. McCaskill’s thigh, then will lead him to source of seizure scent. He will also perform a “sweep” of the room and indicate seizure source if given the command “find seizure.”

As part of standard care in the EMU, the patient is monitored on video EEG throughout the hospitalization. Following clinical events, the attending physician reviews video EEG recording to determine diagnosis and event details. Event descriptions are eventually detailed in a comprehensive EEG report. EEG results were compared to seizure dog response for those cases in which Dr. McCaskill were able to do an assessment. As part of this study, the dog’s response was compared to EEG diagnosis for the clinical event in question, to assess accuracy of the dog’s response.

## 3. Results

### 3.1 Baseline seizure dog survey (pre-interaction)

102 subjects consented to participate in the study and 98 completed the baseline survey before interaction with the seizure dog. Four patients did not complete the entire pre-test survey and were excluded. The median participant age was 36 years (range 18 to 78). 56% indicated they currently own or live with a dog; 15% reported ever having owned a trained service dog. Of the patients who had owned a service dog (n=15), 80% reported that their service dog was trained for seizure response, and the other 20% responded having had service dogs trained for mental health response. 93% (n=91) of participants indicated feeling “extremely comfortable” or “very comfortable” with a service dog being present, and 43% (n=42) indicated that they had previously interacted with a service dog in a medical setting. In terms of training capability, 87% (n=85) of participants believed dogs could be trained to “smell seizures,” 100% believed dogs could “respond to the emotional state of their owner”, and 99% (n=97) believed dogs could “respond to the behavior or actions of their owners.” No subjects reported having safety concerns prior to the interaction with Reggie.

### Post-seizure dog interaction survey

35 of 98 participants interacted with the seizure dog and completed the post-survey. 55 participants completed the pre-survey but did not have the opportunity to interact with the seizure dog due to limitations with Dr. McCaskill’s schedule and availability. Three patients did interact with Reggie but were discharged prior to completing the post-survey. An additional five participants completed both surveys, but the physical post-survey paper form was lost/discarded during the discharge process. 91% (n=32) of participants in the post-survey indicated they felt “ok to have physical contact with the service dog” or had “no preference” regarding physical proximity to the dog. All participants felt there was a potential role for service dogs in medical care, both in hospital and outpatient settings, with 91% responding “yes” to personally experiencing benefits of working with the seizure dog. Free responses regarding benefits of SSD interactions described emotional benefits including “emotional support,” “comfort,” “helped me calm down” “improved my mood” with samples of patient responses highlighted in **Table 1**. No participants reported a negative experience with the dog, nor safety concerns. Regarding seizure detection, after interacting with Reggie, 82% (n=29) of responders responded that he could “definitely” detect seizures, 14% (n=5) said he could “probably” detect seizures, and 3% (n=1) answered “I don’t know.” However, when asked whether respondents believed that Reggie could distinguish between epileptic and non-epileptic seizures, there was less certainty, with 51% (n=18) responding “definitely,” 31% (n=11) “probably” and 17% (n=6) “I don’t know.” No participants reported having any concerns about their personal interactions with our service dog.

**Table 1:**
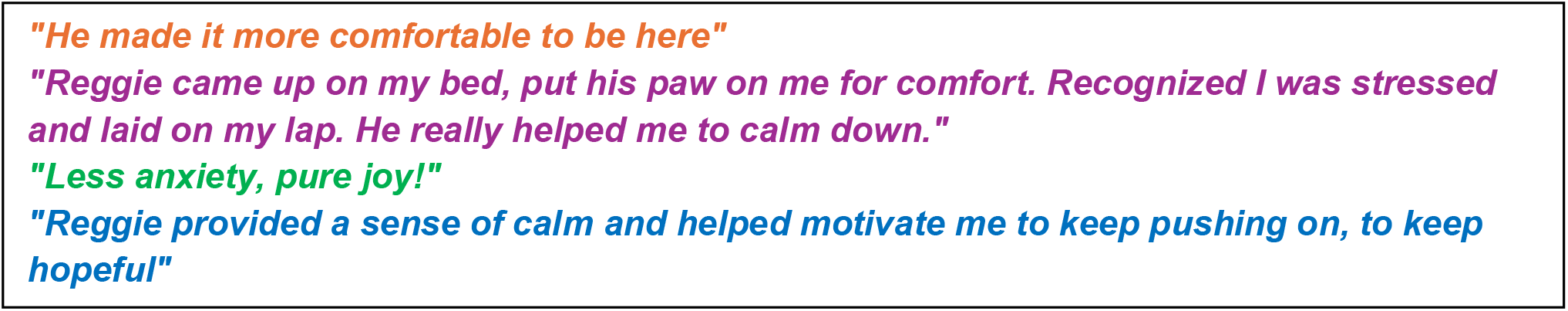
Selection of patient comments about experience with Reggie.

### 3.2 Case examples

**Case 1 –** On day 5 of hospital admission for a stereotactic EEG study, Dr. McCaskill and Reggie are in the hallway of the Epilepsy Monitoring Unit when Reggie indicates seizure alert to Dr. McCaskill and indicates source of seizure as patient’s room. Dr. McCaskill and Reggie enter the room and approximately 1 minute later, the patient begins having a typical clinical seizure; seizure response protocol ensues. After leaving the patient’s room following the seizure, Dr. McCaskill reviews EEG with another epileptologist, confirming electroclinical seizure with right hippocampal onset 56 seconds after Reggie’s initial alert.

**Case 2 -** On rounds on admission day 4 upon neurology team entering the room, Reggie started exhibiting seizure alert behavior spontaneously without command. He indicated the patient was the source of the seizure smell. Review of EEG post rounds confirmed 2 electrographic seizures in that time period, both 17 minutes prior to Reggie’s seizure alert as well as 22 minutes following his alert. Both seizures were focal with impaired consciousness, left temporal onset.

**Case 3** - On day 4 of the patient’s EMU admission, Dr. McCaskill and Reggie were in the patient room for rounds. Reggie spontaneously alerted and indicated the patient as source of a seizure. 13 minutes later, the patient had a focal to bilateral tonic-clonic seizure with the team still in the room, later confirmed on EEG to be epileptic with right temporal ictal onset.

## 4. Discussion

### 4.1 Patient Perspective

Patients were generally eager to participate in this study, and there was a great deal of enthusiasm about incorporating a seizure dog into EMU care. Multiple patients expressed interest in obtaining a personal seizure dog after this experience. In general, the vast majority of participants expressed a high degree of comfort with the presence of a seizure dog both before and after having interactions with Reggie. No patients reported safety concerns before or after interacting with the dog, and many patients reported specific benefits after interacting with Reggie, particularly having to do with emotional comfort.

Our findings are in line with prior research that emphasizes the potential for service dogs to improve emotional well-being and the overall experience of patients and caregivers. Ours is the first survey study to examine patient attitudes toward integration of service dogs unknown to the patient in an inpatient setting specifically. Prior survey studies of people with epilepsy who owned dogs showed that owners universally reported psychological benefits to seizure dog ownership including companionship and reduced stress levels.^4,5^ Seizure dog ownership may also reduce caregiver burden, which has been shown to be significant with impact on quality of life for both patients and caregivers.^10^ Strong et al. (2002) reported a 43% reduction in convulsive seizures among patients using seizure alert dogs, attributing improvement to reduced stress due to the dogs’ presence. In addition to clinical benefits, the same study also found that patients reported reduced anxiety and greater independence, with no adverse effects recorded.^11^ Given the initial data from this study indicating high degree of enthusiasm for SSD integration into inpatient care with no reported safety concerns and potential for improvement of patient well-being, we hope that this study will inspire future inpatient studies integrating SSD into the EMU setting.

### 4.2 Lessons Learned about Seizure Detection

In addition to patients’ comfort and enthusiasm for seizure dogs, many patients also expressed strong beliefs about the seizure detection capabilities of these dogs. Approximately 85% indicated strong confidence that dogs could be trained to detect seizures, with many citing anecdotal evidence or personal experiences with service animals. Our case examples offer additional evidence that trained seizure dogs may have the capacity to recognize epileptic seizures in the inpatient setting. Seizure dogs have the potential to augment epilepsy care through seizure recognition and response. Patients are often unaware of seizures, with one EMU study finding that only 26% of patients were always aware of their seizures, and 30% were never aware of their seizures. Seizure dogs could potentially help increase seizure awareness in select patients with poor self-awareness of seizures.^12^

Scientific validation of this ‘seizure-sniffing’ ability remains limited to case reports and case series, but recent research confirms that some dogs can detect seizures by scent. Studies utilizing data from epilepsy monitoring units (EMUs) have faced challenges, particularly when psychogenic nonepileptic seizures complicate the data. For example, one study compared two patients’ seizure dogs’ responses as compared to EEG found that one seizure dog failed to identify most seizures and the other dog identified the patient’s events accurately, but the events were confirmed to be non-epileptic.^13^

### 4.3 Limitations and Challenges

There are limitations to our study. There may be an element of response bias because the patients who chose to participate likely had a priori interest in dogs and this enthusiasm is reflected in their survey responses. There may also be some bias in our case examples given the retrospective observational non-randomized study design. Previous studies on seizure dogs have suggested that dogs may respond to handlers’ behavioral cues as well as responding to trained scent recognition.^14^ Although Reggie’s handler was not aware of EEG findings at the moments when Reggie spontaneously signaled, Dr. McCaskill may have given nonverbal cues based on clinical judgement that impacted Reggie’s response. There is anecdotal evidence from our case examples that Reggie can spontaneously identify epileptic events, but the encounters were infrequent enough that this was difficult to test systematically. A prior EMU case series suggested that seizure dogs may respond to non-epileptic events, but this prior study involved patients’ own service animals.^13^ It is logistically difficult to design a study in an inpatient clinical setting where a hospital seizure dog can have seizure detection tested in a systematic way, given that the dog cannot be present 24 hours per day, many seizures occur overnight, and there is an element of chance as to when the dog’s presence will overlap with patients’ spontaneous seizures.

Our study demonstrates that seizure dogs can be successfully integrated into an EMU setting; however, the prospect of integrating seizure dogs into patient care on a wider scale presents many logistical challenges. These challenges fall into two main categories: the training and cost of seizure dogs, and the practical feasibility of introducing these animals into medical environments. Medical facilities often have strict regulations regarding animal presence, which can complicate the integration of service dogs. Additionally, there may be concerns about hygiene, patient safety, and potential allergies among other patients and staff.^15^ Our survey results reassured us that these concerns may be overstated as long as patients are amenable to dog interactions and the seizure dogs are well-trained.

Training dogs to recognize seizure-related scents or behaviors can take months or even years, depending on the dog and the program. The hospital setting requires a specific set of training and skills to maintain appropriate behavior with all of the distractions present in the hospital environment. Our particular SSD had extensive training as previously detailed. Not all dogs are the same in terms of service potential and seizure recognition potential, and so far, there are no established guidelines for ideal breeds for training or ideal training.^16,17,18^ A survey study of people with epilepsy who owned dogs in France showed that spontaneous presence or absence of alerting behaviors had less to do with dog breed and more to do with dog personality and dog-owner relationship.^19^ Training for seizure dogs can be costly and time intensive.^20^ This high cost and high effort can limit accessibility for patients who may benefit from seizure detection dogs. There are charitable organizations that fund and train seizure dogs for select recipients.^21^

### 4.4 Future Directions

Most seizure dog studies, including the randomized controlled EPISODE trial, have been limited by small sample sizes and short follow-up periods.^8^ Longer-term studies with larger sample sizes could provide more robust data on the sustained effectiveness of seizure dogs. Additional data from EMU settings could help determine whether seizure dogs may augment the patient experience and potentially help with seizure alert and response in select cases. While it remains unclear to what extent most seizure dogs have the potential to differentiate between epileptic and nonepileptic seizures, they have the potential to provide comfort and event awareness in either event type. There is also a need to help refine guidelines for seizure dog training protocols and explore ways to implement seizure dogs into patient care in a cost and time efficient manner.

## 5. Conclusions

Seizure dogs offer multifaceted potential benefits to the care of patients with epilepsy, including companionship, comfort, safety, seizure recognition, and response. There is widespread enthusiasm among patients for seizure dogs, and our results demonstrate that this enthusiasm translates to the inpatient EMU setting as well as outpatient setting. It is feasible to safely integrate a trained seizure dog into the EMU, and seizure dogs may offer additional insights into seizure awareness that complement and augment the inpatient EMU experience.

## Supporting information

post-test survey

pre-test survey

## Data Availability

All data produced in the present study are available upon reasonable request to the authors.

## Data Availability Statement

De-identified data may be available from the corresponding author upon reasonable request and with approval from the Oregon Health & Science University Institutional Review Board.

