## Supplementary material for "Patient Perceptions of a Seizure Service Dog in the Epilepsy Monitoring Unit": post-test survey

### EMU-STUDY SURVEY

| | |
|---|---|
| **1.** Participant Number: | **2.** Age: |

| 3. How comfortable did you feel with a service dog being present during your stay in the hospital/EMU (or time in the clinic)? | | | | | |
|---|---|---|---|---|---|
| Extremely comfortable ☐ | Very comfortable ☐ | Somewhat comfortable ☐ | Somewhat uncomfortable ☐ | Very uncomfortable ☐ | Extremely uncomfortable ☐ |

| 4. How much did you like the seizure dog? | | | | | | |
|---|---|---|---|---|---|---|
| Loved the seizure dog ☐ | Liked a lot ☐ | Liked a little ☐ | Dislike them a little ☐ | Dislike them a lot ☐ | Hated him ☐ | Don't have strong feelings about them either way ☐ |

| 5. Do you think that the EMU/epilepsy clinic service dog can detect seizures? | | | | |
|---|---|---|---|---|
| Yes-Definitely ☐ | Yes-Probably ☐ | I don't know-it's possible ☐ | No- Probably not ☐ | No-definitely not ☐ |

| 6. Do you think that the EMU/epilepsy clinic service dog can tell the difference between epileptic seizures and non-epileptic seizures (ie spells that look like seizures, but do not show evidence of seizure activity on the EEG)? | | | | |
|---|---|---|---|---|
| Yes-Definitely ☐ | Yes-Probably ☐ | I don't know-it's possible ☐ | No- Probably not ☐ | No-definitely not ☐ |

| 7. Do you think there is a role for service dogs in the hospital / EMU? | | | | |
|---|---|---|---|---|
| Yes - definitely, they can help patients ☐ | Yes - probably, they can help some patients ☐ | I don't know ☐ | No - they don't help Patients ☐ | No - they might harm patients ☐ |

| 8. Do you think there is a role for service dogs in outpatient seizure clinic? | | | | |
|---|---|---|---|---|
| Yes - definitely, they can help patients ☐ | Yes - probably, they can help some patients ☐ | I don't know ☐ | No - they don't help Patients ☐ | No - they might harm patients ☐ |

9. At what distance should the service dog be from you in the EMU to make you most comfortable?

| At least 6 feet away | at least 3 feet away | at bedside | ok with dog touching you | No preference |
|---|---|---|---|---|
| ☐ | ☐ | ☐ | ☐ | ☐ |

| 10. Did you personally experience any benefits of working with the service dog in the EMU/seizure clinic? | Yes ☐ No ☐ |
|---|---|
| If Yes, please describe: | |

| 11. Were there any downsides of working with the service dog in the EMU/seizure clinic? | Yes ☐ No ☐ |
|---|---|
| If Yes, please describe: | |

| 12. Did you have any safety concerns about working with the service dog in the EMU/seizure clinic? | Yes ☐ No ☐ |
|---|---|
| If Yes, please describe: | |

13. Do you have any questions or concerns about participating in this study? Anything else you would like to share?
